# Antigenic minimalism of SARS-CoV-2 is linked to surges in COVID-19 community transmission and vaccine breakthrough infections

**DOI:** 10.1101/2021.05.23.21257668

**Authors:** A.J. Venkatakrishnan, Praveen Anand, Patrick Lenehan, Pritha Ghosh, Rohit Suratekar, Abhishek Siroha, Dibyendu Roy Chowdhury, John C. O’Horo, Joseph D. Yao, Bobbi S. Pritt, Andrew Norgan, Ryan T. Hurt, Andrew D. Badley, John D. Halamka, Venky Soundararajan

**Affiliations:** nference, Cambridge, Massachusetts 02139, USA; nference Labs, Bengaluru, Karnataka, India; Mayo Clinic, Rochester, Minnesota 55902, USA

**Author notes:** Correspondence to: Venky Soundararajan, A.J. Venkatakrishnan. Equal first authors (A.J. Venkatakrishnan, Praveen Anand).

## Abstract

The raging COVID-19 pandemic in India and reports of “vaccine breakthrough infections” globally have raised alarm mandating the characterization of the immuno-evasive features of SARS-CoV-2. Here, we systematically analyzed 1.57 million SARS-CoV-2 genomes from 187 countries/territories and performed whole-genome viral sequencing from 53 COVID-19 patients, including 20 vaccine breakthrough infections. We identified 89 Spike protein mutations that increased in prevalence during at least one surge in SARS-CoV-2 test positivity in any country over a three-month window. Deletions in the Spike protein N-terminal domain (NTD) are highly enriched for these ‘*surge-associated mutations*’ (Odds Ratio = 41.8, 95% CI: 6.36-1758, p-value = 7.7e-05). In the recent COVID-19 surge in India, an NTD deletion (ΔF157/R158) increased over 10-fold in prevalence from February 2021 (1.1%) to April 2021 (15%). During the recent surge in Chile, an NTD deletion (Δ246-253) increased rapidly over 30-fold in prevalence from January 2021 (0.86%) to April 2021 (33%). Strikingly, these simultaneously emerging deletions associated with surges in different parts of the world both occur at an antigenic supersite that is targeted by neutralizing antibodies. Finally, we generated clinically annotated SARS-CoV-2 whole genome sequences and identified deletions within this NTD antigenic supersite in a patient with vaccine breakthrough infection (Δ156-164) and other deletions from unvaccinated severe COVID-19 patients that could represent emerging deletion-prone regions. Overall, the expanding repertoire of Spike protein deletions throughout the pandemic and their association with case surges and vaccine breakthrough infections point to antigenic minimalism as an emerging evolutionary strategy for SARS-CoV-2 to evade immune responses. This study highlights the urgent need to sequence SARS-CoV-2 genomes at a larger scale globally and to mandate a public health policy for transparent reporting of relevant clinical annotations (e.g. vaccination status) in order to aid the development of comprehensive therapeutic strategies.

## Introduction

The ongoing COVID-19 pandemic has infected around 160 million people and killed more than 3 million people worldwide, as of May 2021^1^. The continual emergence of SARS-CoV-2 variants with increased transmissibility and capacity for immune escape, such as B.1.1.7 (“UK variant”) and P.1 (“Brazilian variant”), threatens to prolong the pandemic through devastating outbreaks such as the one currently being witnessed in India^2^. While multiple vaccines have demonstrated high effectiveness in clinical trials and real world studies^3–5^, there have been reports of “vaccine breakthrough infections” with SARS-CoV-2 variants^6,7^. A recent study described two such cases in New York, at least one of which occurred despite confirmation of a robust neutralizing antibody response. Variant classification schemes have been developed by the US Centers for Disease Control and Prevention (CDC)^8^ and the World Health Organisation (WHO)^9^ based on factors such as prevalence, evidence of transmissibility and disease severity, and ability to be neutralized by existing therapeutics or sera from vaccinated patients. Early and rapid detection of these emerging Variants of Concern/Interest is imperative to combat and contain the ongoing pandemic and future outbreaks.

Mapping the mutational landscape of SARS-CoV-2 in the context of natural and vaccine-induced immune responses is critical to understand the virus’s molecular strategies for immune evasion. To this end, neutralizing antibodies which target the receptor-binding domain (RBD) or the N-terminal domain (NTD) of the Spike protein have been isolated from the sera of COVID-19 patients^10–12^. Recent studies contemporaneously found that several neutralizing antibodies target a single antigenic supersite in the NTD of the Spike protein^13,14^. The NTD is also a hotspot for in-frame deletions in the SARS-CoV-2 genome, with four recurrent deletion regions (RDRs) identified^15^. Several such deletions have been experimentally demonstrated to reduce neutralization by NTD-targeting neutralizing antibodies^13,15^. Whether additional deletions are emerging in SARS-CoV-2 variants that drive case surges or vaccine breakthrough infections needs to be determined.

Since the beginning of the pandemic concerted global data sharing efforts have led to the rapid development of large-scale genomic and epidemiological COVID-19 resources. 1.57 million SARS-CoV-2 genomes from 187 countries/territories have been deposited in the GISAID database^17^ (**Figure 1**). In addition, we performed whole-genome sequencing of SARS-CoV-2 from patients at the Mayo Clinic that had recently developed SARS-CoV-2 infections or “vaccination breakthrough” infections. On the epidemiology front, population-level metrics including SARS-CoV-2 test positivity rates are being collected from 219 countries in databases such as OWID^16^. Such unprecedented availability of genomic-epidemiological data combined with patient-level clinical genomic data provides a timely opportunity to systematically characterize the immuno-evasive features of SARS-CoV-2.

**Figure 1.**
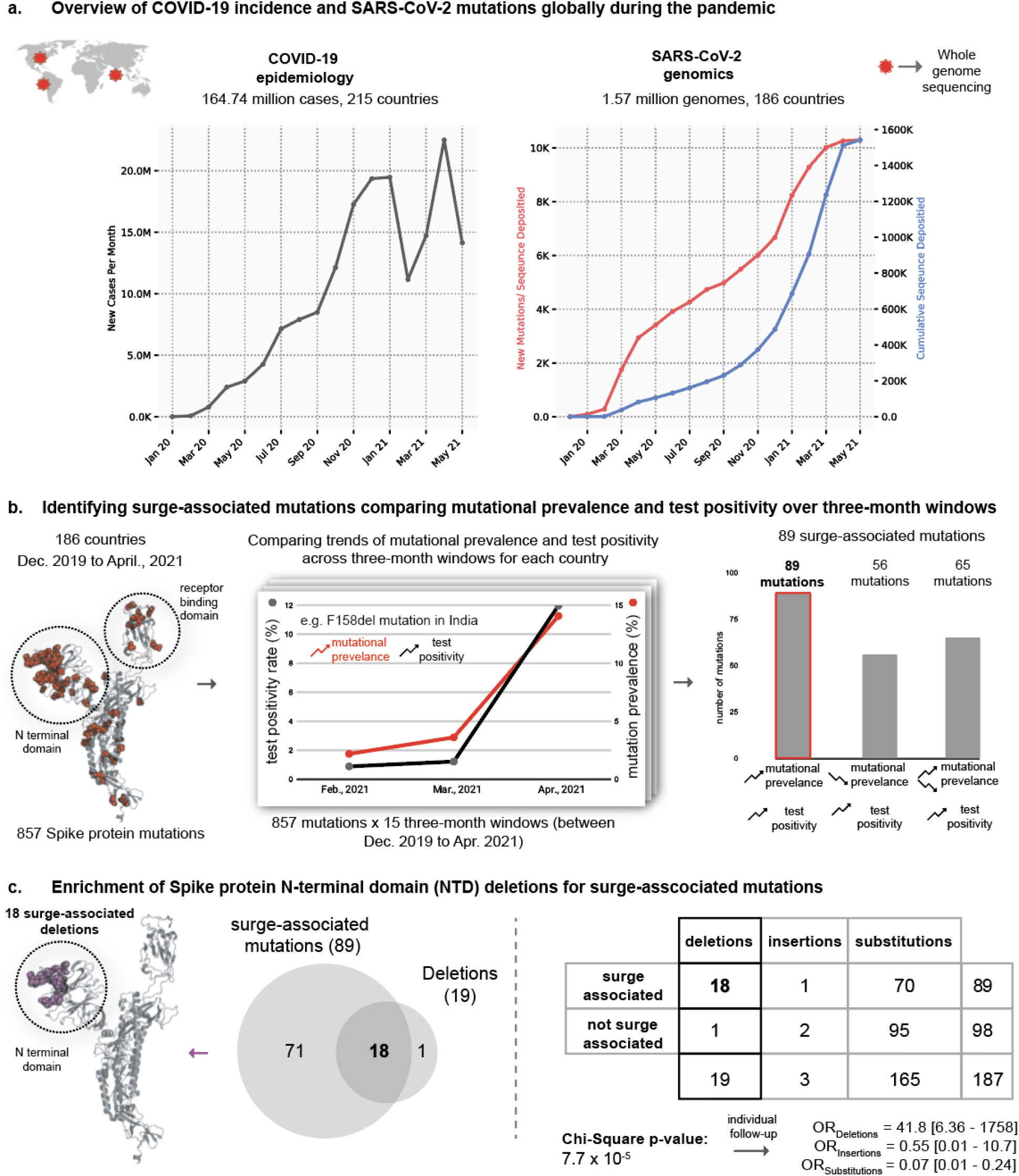
Identifying potential SARS-CoV-2 variants contributing to COVID-19 surge. (a) Overview of COVID-19 prevalence and SARS-CoV-2 variants globally during the pandemic. (b) Identifying surge-associated mutations comparing mutational prevalence and test-positivity over three-month windows. Structure shows the sites of all the 857 mutations (as red spheres) on a subunit of the Spike protein — each site can harbor multiple mutations. Chart shows correlation between trends of mutational prevalence (e.g. F158del) and test positivity during a three-month window (e.g. Feb, 2021 to Apr. 2021 in India). These correlations are computed for all three-months across all the countries being studied. Barchart shows the frequency of mutations that fall in each combination of trends: (i) 89 surge-associated mutations: mutational prevalence and test positivity are increasing monotonically, (ii) 56 mutations where mutational prevalence is increasing monotonically and test positivity decreasing monotonically, and (iii) 65 mutations where test positivity is increasing monotonically and the trend in mutational prevalence is mixed. Other categories (e.g. test positivity decreasing) are not shown for clarity. (c) The enrichment of N-terminal domain deletions among surge-associated mutations. Structure shows the sites of all the 18 surge-associated deletion mutations (as magenta spheres) on a subunit of the Spike protein. Venn diagram shows the overlap of surge-associated mutations and deletion mutations. The 2×3 table shows the distribution of mutations based on surge-association and mutation category (deletion, insertion, substitution). Chi-square test results are also shown.

In this study, we uncover that deletion mutations in the Spike protein have a high likelihood of being associated with surges in community transmission. Further, we identify that rapidly surging SARS-CoV-2 variants in India and Chile have acquired new deletions in an antigenic supersite enabling evasion of neutralizing antibodies. Based on a global analysis of deletions we also highlight that the repertoire of deletion-prone regions of the Spike protein is expanding during the course of the pandemic pointing to an evolutionary strategy of “antigenic minimalism” to evade immune responses. Finally, using whole genome sequencing linked to clinical annotations derived from electronic health records, we also identify deletion mutations in SARS-CoV-2 from COVID-19 patients with infection/vaccine-breakthrough infections, also mapping near the antibody-binding site and thus representing candidates for vaccine escape mutations.

## Results

### Deletions are enriched for association with surges in community transmission of SARS-CoV-2

Analysis of 1,567,718 SARS-CoV-2 genome sequences obtained from the GISAID database^17^(**Figure 1a**) revealed the presence of 857 amino acid mutations (missense, insertions and deletions) in the Spike protein. This list included mutations that were observed in at least 100 SARS-CoV-2 genome sequences in order to exclude random occurrences from sequencing errors. The list of 857 mutations included 816 substitutions (95.2%), 37 deletions (4.3%), and 4 insertions (0.4%). To identify the mutations associated with surges in the community transmission of COVID-19 (“surge-associated mutations’’), we shortlisted mutations that increased in prevalence monotonically during periods of monotonically increasing test positivity (**Figure 1b**). We identified 89 mutations that increased in prevalence during one or more surges in test positivity rate (with minimum change in test positivity rate >= 5% and minimum change in mutational prevalence >=5% over the three-month window), in any country, over a three-month time interval. This approach recapitulated 39 out of 56 (70%) mutations known to be present in the CDC variants of interest or concern, including D614G, E484K, N501Y, P681H, P681R, ΔH69/V70, and ΔY144 (**Figure S1**).

Further, we investigated whether a class of mutations (missense, insertions and/or deletions) is enriched for association with surges. We applied a stringent threshold for inferring the trends by ensuring that mutational prevalence and positivity data is present over the complete time-window of three months. This reduced the number of mutations from 857 to 255. Interestingly, we observed an enrichment of deletion mutation in the surge with 18 out of 19 deletions associated with the surge (**Figure 1c**; Chi-square Test p-value = 7.7e-05; Odds Ratio = 41.8, 95% CI: 6.36-1758). These surge-associated deletions in the Spike protein occur exclusively in the N-terminal domain, which is interesting in light of the recently identified recurrent deletion regions (RDRs) in the N-terminal domain^15^. This raises the possibility that SARS-CoV-2 may be acquiring deletion mutations to evolve new variants that drive the surges in community transmission of COVID-19.

### Surging SARS-CoV-2 variants in India and Chile have acquired new deletions in an antigenic supersite enabling evasion of neutralizing antibodies

Recently there have been massive surges of COVID-19 community transmission in a few countries around the world, but most prominently in India^18^ and Chile^19,20^. In order to identify the mutations that are associated with these surges, we systematically analysed the mutations that increased monotonically in prevalence correlated with the monotonic increase in test positivity between February and April 2021 (**Figure 2a** and **Figure 3a**).

**Figure 2.**
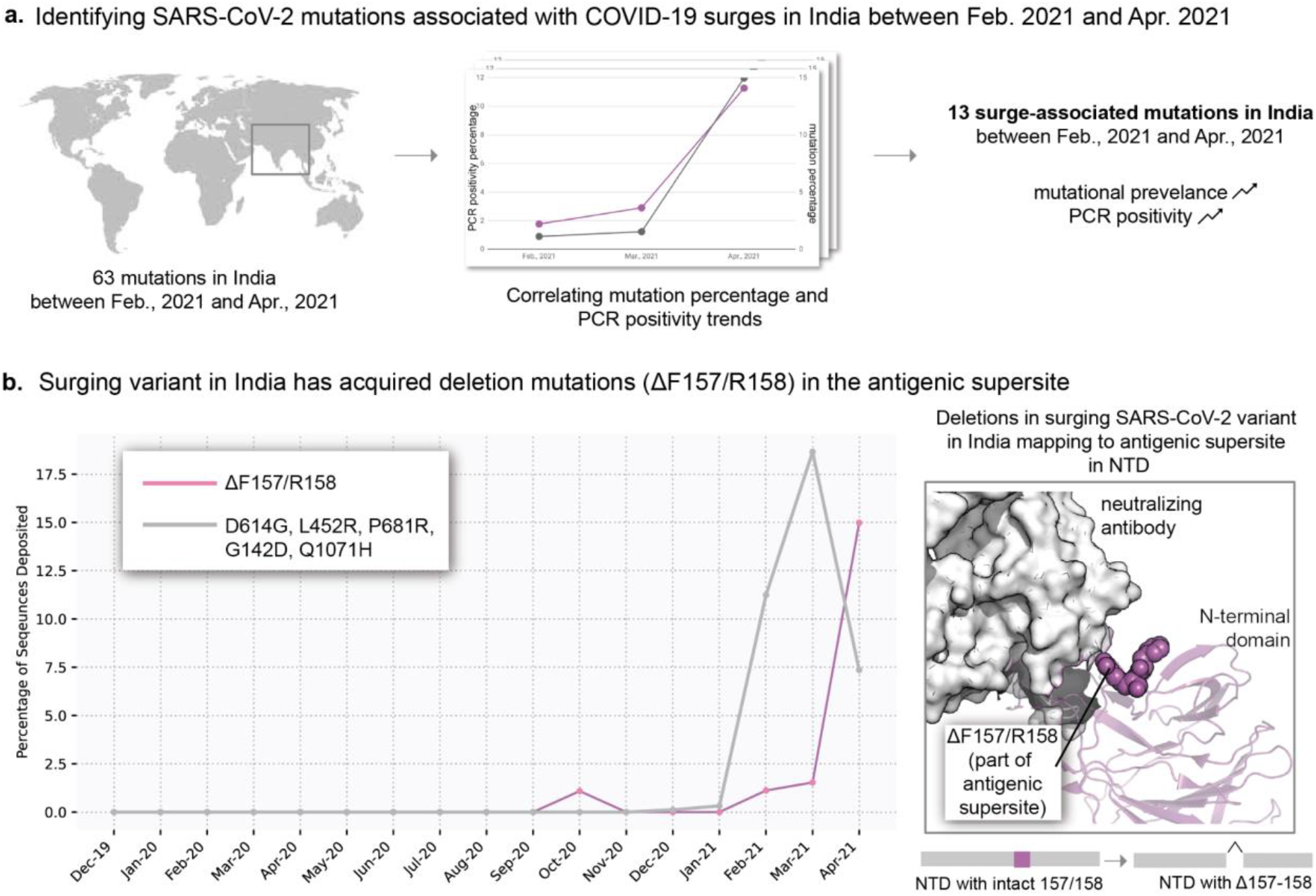
Surging SARS-CoV-2 variant in India has acquired deletions in an antigenic supersite of the Spike protein N-terminal domain. (a) Identification of mutations in the SARS-CoV-2 Spike protein that are associated with the COVID-19 surge in India between February and April 2021, based on correlations between mutational prevalence and test positivity. 13 mutations were found to be positively correlated with the surge over the three-month window. (b) Tracking the prevalence of ΔF157/R158 (magenta) in the surging variant in India compared to other prevalent mutations (gray). The inset shows the location of these two residues on the Spike protein structure.

**Figure 3.**
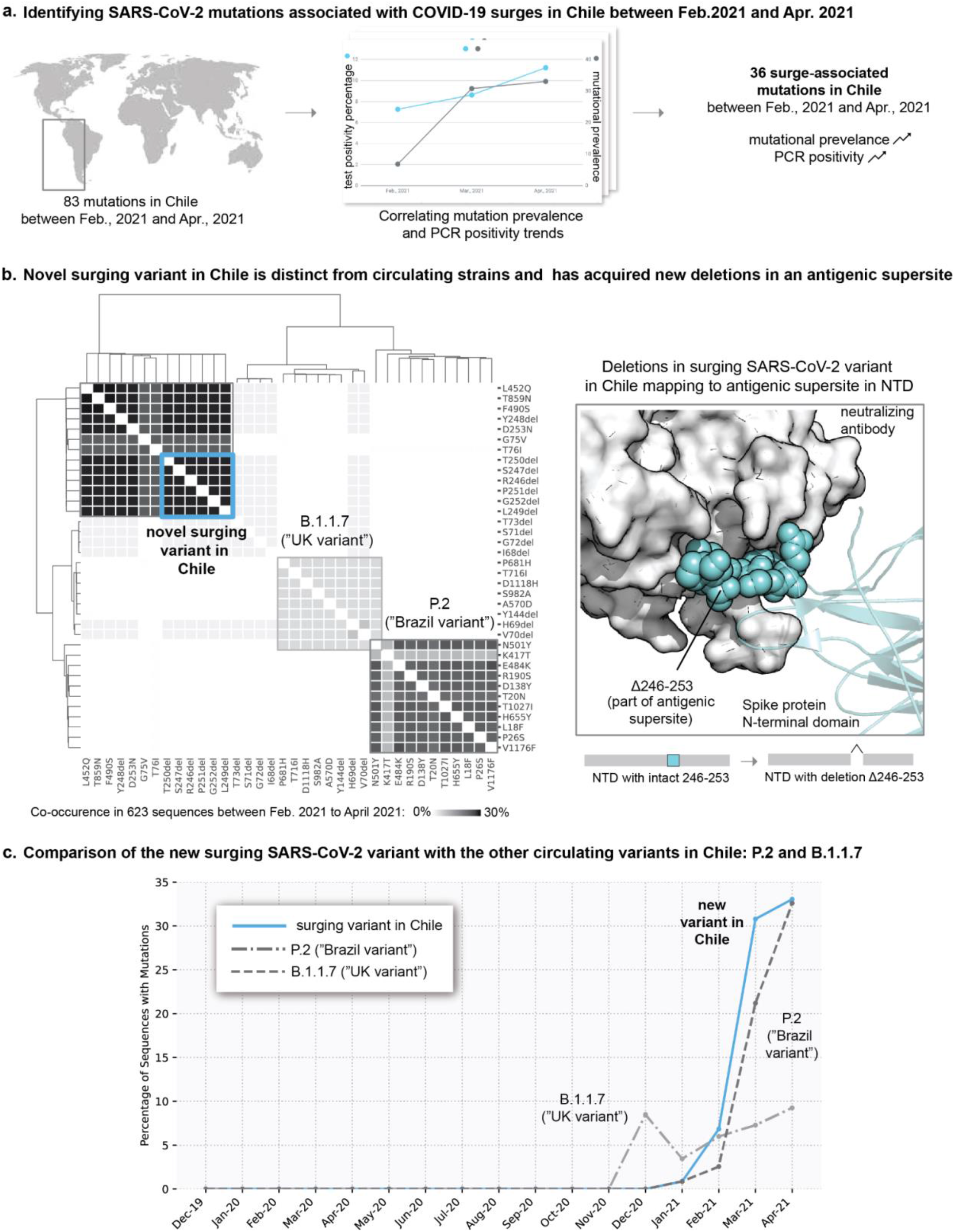
New surging variant in Chile has acquired deletions in the antigenic supersite of the Spike protein N-terminal domain. (a) Identification of mutations in the SARS-CoV-2 Spike protein that are associated with the COVID-19 surge in Chile between February and April 2021, based on correlations between mutational prevalence and test positivity. A total of 36 mutations were found to be increasing monotonically in prevalence with the surge over the three-month window. (b) The clustering of these mutations based on their co-occurrence (number of sequences with both mutations present) using the complete method (farthest point algorithm) reveals three dominant clusters. The clusters highlight that the novel surging variant in Chile is distinct from the circulating variants. (c) Rapid increase of the prevalence of the deletion mutations in the surging variant in Chile (blue) compared to the mutations in the circulating variants (B.1.1.7 and P.2).

In India, 13 Spike protein mutations were correlated with the massive surge (“second wave of infections’’) in April 2021 (**Table S1**). The most prevalent mutations included D616G, L452R, P681R, G142D and Q1071H (**Figure 2b** and **Figure S2**). Interestingly, there is also a rapidly emerging deletion in the NTD (ΔF157/R158) that has increased 13.6 fold in prevalence from 1.1% (of 1254 sequences) in February 2021 to 15% (of 367 sequences) in April 2021. F157 and R158 reside in the antigenic supersite, which is recognized by a number of NTD-targeting neutralizing antibodies^14,21^ (**Figure 2b**). Importantly, owing to the recent emergence of this deletion it had not been identified at the time of the prior characterization of Spike protein deletions^15^, and thus we suggest that ΔF157/R158 represents a novel distinct fifth recurrent deletion region.

In Chile, 36 Spike protein mutations are correlated with the surge in April 2021 (**Figure 3b, Table S2**). Clustering these mutations by co-occurrences in sequences shows the emergence of a new variant characterized by a novel in-frame deletion resulting in the loss of residues 246-253 and a D253G substitution (Δ246-253), which has increased 38.4 fold in prevalence from January to April 2020 (0.86% to 33.0%) (**Figure 3b**). This new surging variant in Chile is independent of two other concurrently circulating variants: P.2 (first identified in Brazil) and B.1.1.7 (first identified in the UK) (**Figure 3b**). Interestingly, residues 246-253 are part of the “supersite loop” in the same NTD antigenic supersite that includes F157 and R158 (**Figure 3b**)^14,21^.

Taken together, this analysis of India and Chile highlights two simultaneously surging strains in different parts of the world that have both acquired deletions in the NTD antigenic supersite (ΔF157/R158 in India, Δ246-253 in Chile) that is highly targeted by neutralizing antibodies. Indeed, deletions in the NTD domain have been previously shown to diminish the binding of neutralizing antibodies^13,15^. This suggests that the surging SARS-CoV-2 variants in India and Chile may have acquired NTD deletions in the antigenic supersite in order to evade neutralizing antibodies and achieve immune escape. Indeed, mutations in this region (at P251, G252, and D253) were also found in neutralization escape mutants *in vitro*^*13*^. From a viral evolution standpoint, these observations raise the question of whether SARS-CoV-2 is expanding its repertoire of deletable regions in the Spike protein as the pandemic progresses.

### Recurrent deletion regions in the Spike protein emerge and expand over the course of the pandemic

In order to understand whether the deletable regions in the Spike protein are increasing, we examined the distribution of deletion frequencies for all amino acids in the Spike protein sequence from over 1.3 million sequences as of April 30, 2021(**Figure 4a**,**b**; see **Methods**). This analysis includes more than 10 fold the number of sequences compared to the previous analysis of recurrent deletion regions that was based on 146,795 sequences as of October 24, 2020 (**Figure 4a**; see **Methods**).

**Figure 4.**
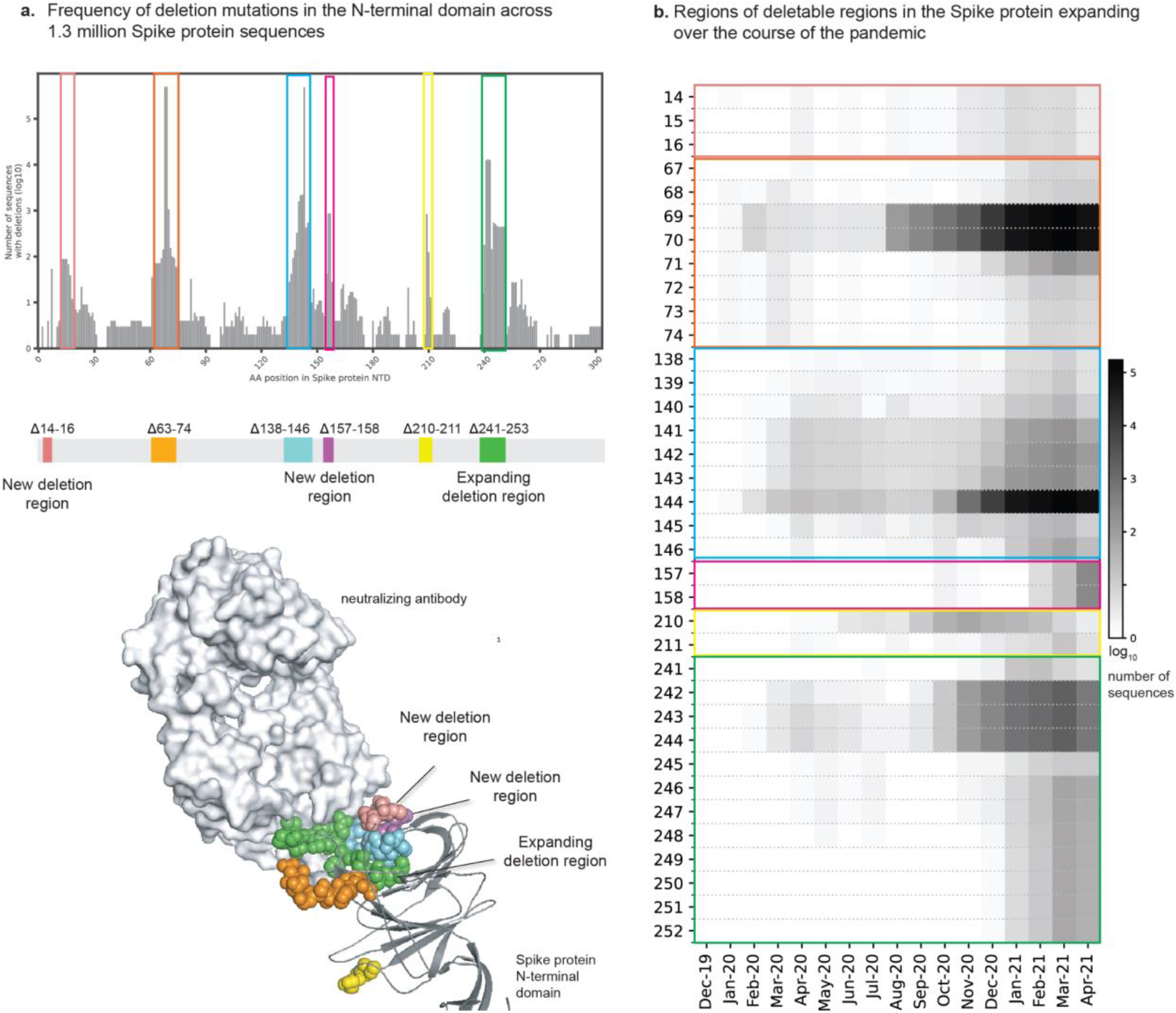
The repertoire of deletions in the Spike protein N-terminal domain is expanding over the course of the pandemic. (a) Frequency of occurrences of deletion mutations in the N-terminal domain across 1.3 million Spike protein sequences (as of April 30, 2021). The recurrent deletion regions, both known as well as new, are illustrated schematically and mapped on the structure of the Spike protein. Positions corresponding to the deletion mutations in the Spike protein are shown as colored spheres and the neutralizing antibody is shown using a surface representation in grey (b) Heatmap showing the expansion of “deletable” regions in the course of the pandemic, where the rows denote residue positions in the Spike protein and columns denote the time course of the pandemic (in months). The boxes denote the frequency of the deletion mutation across the world in that month. The color of the boxes corresponds to a frequency of 1 to 100,000 sequences shown on a log10 scale. These deletion mutations are shown on the 3D structure of the Spike protein. Positions corresponding to deletion mutations are shown as spheres.

In addition to ΔF157/R158 (the new surge-associated deletion in India; **Figure 2**), we observed that residues 14-18 (QCVNL) are deleted more frequently than expected based on the background distribution (see **Methods**). Interestingly, these residues are part of the N-terminal region of the same antigenic supersite (**Figure 4a**), and mutations in this region (at C15 and L18) were common among neutralization escape mutants^13,14^. Furthermore, this deletion region is emerging recently — most viral genomes containing one or more deletions in this region were deposited after October, 2020^15^. We also identified potential recurring deletions at residues S640/N641 and 675-681 (QTQTNSP), the latter of which directly precedes the Spike protein furin cleavage site that we and others have described previously^22–25^. It is notable that these are the only recurring deletion regions observed to date that are outside of the NTD (and thus outside of the antigenic supersite).

We also recognized that recurrent deletion regions appear to have the capacity to expand (i.e., to involve more flanking amino acids) over time. For example, the Δ246-253 deletion in one of the surge associated Chile variants can be viewed as an expansion of the previously defined recurrent deletion region RDR4 (Δ242-248)^15^ (**Figure 5b**). This expansion is likely of high immunologic relevance, as not only do these residues map to the supersite loop, but mutations in this region (at P251, G252, and D253) were also found in neutralization escape mutants *in vitro*^*13*^. Further biochemical experiments can determine whether these previously uncharacterized NTD deletions (residues 14-18, 156-159, and 249-253) impact the binding of NTD-targeted neutralizing antibodies or the capacity of sera from vaccinated individuals to neutralize the virus.

**Figure 5.**
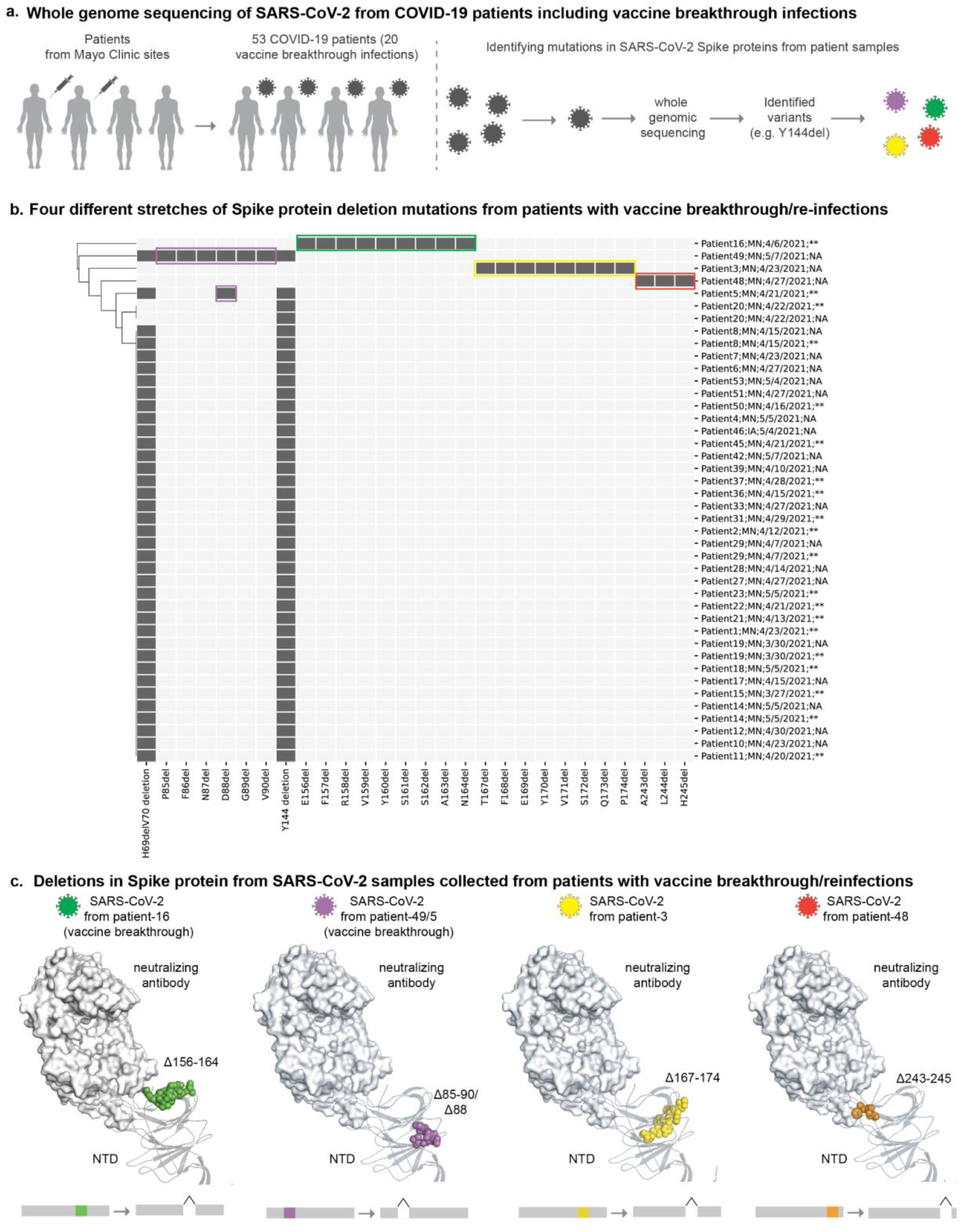
Deletion mutations present in the Spike proteins sequences derived from clinically annotated COVID-19 patients. (a) Whole genome sequencing of SARS-CoV-2 from COVID vaccination breakthrough/reinfections from Mayo Clinic. (b) Four different stretches of Spike protein deletions from the patients with vaccine breakthrough or reinfections of COVID-19. In the heatmap, rows denote patients (state, date of sample and vaccination status are shown) and columns denote deletion mutations. ** indicates vaccinated individuals and ‘NA; denotes suspected re-infection. Filled boxes denote the presence of deletion mutations, which are shown on the 3D structure of the Spike protein in panel c. (c) Positions corresponding to deletion mutations are shown as spheres.

Taken together, our analysis highlights both the emergence of novel deletion regions and the expansion of previously defined deletion regions over the past several months. These should be monitored as candidates for emerging hotspots of deletion.

### Whole-genome sequencing of SARS-CoV-2 genomes from COVID-19 patients with vaccine breakthrough reveals the presence of distinct deletions in the N-terminal domain

The genomic-epidemiology analysis presented above based on publicly accessible data has shown that SARS-CoV-2 appears to be acquiring deletion mutations to evade neutralizing antibodies and that the deletable regions are expanding over the course of the pandemic. However, the genome sequences deposited in publicly accessible databases (e.g. GISAID) lack any clinical or phenotypic annotations such as vaccination status or disease severity of the linked COVID-19 patients. To address this, we performed whole genome viral sequencing from 53 COVID-19 patients from the Mayo Clinic health system, for whom we have complete longitudinal health records and vaccination history. Of these, 20 cases were vaccine breakthrough infections, with the infected individuals having been fully vaccinated at the time of their positive SARS-CoV-2 test. In total, we identified 92 unique mutations, of which 28 are deletions (**Figure 5a**). All observed Spike protein deletions in this cohort occurred in the NTD, with Δ144 and ΔH69/V70 showing the highest prevalence (64% and 62%, respectively).

Interestingly, we identified four Spike protein variants harboring one or more deletions that warranted follow up (**Figure 5b**). Two of these novel deletions (Δ156-164, ΔD88) were isolated from previously vaccinated individuals. Whether the deletions were already present at the times of infection or evolved within these individuals under the pressure of vaccine-induced immunity is not known.

One patient who had received two doses of the Pfizer vaccine BTN162b2 in January 2021 was subsequently infected in April. The virus recovered from this patient contained a Δ156-164 deletion (**Figure 5b**). Given the prominent ΔF157/R158 deletion in India, this corroborates our prior observation that recurrent deletion regions can expand and suggests that ΔF157/R158 may actually be part of a larger deletion-prone region. More importantly, the possibility that this variant with a large contiguous deletion within the antigenic supersite was able to infect a fully vaccinated individual mandates further characterization of the potential immuno-evasive effects of deletions in this region.

In another breakthrough infection (Ad26.COV2.S), a patient who had received the J&J vaccine in early April 2021 subsequently tested positive for SARS-CoV-2 by the end of the month. The recovered viral genome harbored a ΔD88 deletion in addition to ΔH69/V70 and ΔY144 (**Figure 5b**). This ΔD88 deletion is quite rare, having been detected in only five of the 1.3 million total sequences deposited (as of April 30, 2021). That said, there was one unvaccinated individual in this Mayo Clinic cohort who experienced severe COVID-19 with a virus containing a ΔD85-90 deletion (**Figure 5b**). There are three sequences in the GISAID database containing this full stretch deletion, two of which are from Germany and one from Texas (**Table S3**). While still too rare to infer its future trajectory, deletions in this region should be epidemiologically monitored in the coming months, particularly to determine whether they impact vaccine effectiveness.

Finally, the viral genome recovered from another severe COVID-19 case in an unvaccinated individual contained a Δ167-174 deletion (**Figure 4b**). While these residues are not within the antigenic supersite itself, they do reside in a loop proximal to it, the deletion of which would almost certainly impact the three-dimensional structure of the supersite (**Figure 4c**). Interestingly, residues 168-172 appear to be a potential emerging deletion hotspot in the GISAID dataset, only narrowly missing classification as a recurring deletion region based on our defined criteria (see **Methods**). Thus, this is a candidate region to emerge as a bonafide recurring deletion region in the near future.

This real-time genomic surveillance of SARS-CoV-2 genomes from COVID-19 patients with linked phenotypic and clinical annotations greatly complements our analysis of the publicly accessible unannotated sequences. Specifically, we have identified deletions within the NTD antigenic supersite which are associated with vaccine breakthrough infections (e.g., Δ156-164), and we have identified other deletions in SARS-CoV-2 isolated from unvaccinated severe COVID-19 patients that could represent emerging recurring deletion regions (e.g., Δ167-174). Taken in the context of previous structural and experimental studies, these deletion patterns seem to represent a historical footprint and future trajectory of the antigenic minimalism strategy employed by SARS-SoV-2 to evade natural and vaccine-induced immunity.

## Discussion

The worldwide mass vaccination campaign has had a profound impact on COVID-19 transmission. However, certain variants are less susceptible to neutralization by sera from vaccinated individuals and convalescent COVID-19 patients^26,27^. Such findings motivate the need to vigilantly track the emergence of new variants and to determine whether they are likely to cause surges or vaccine breakthrough infections. Here, through an integrated analysis of genomic-epidemiology and clinical genomics, we found that (i) deletions are strongly associated with surges in community transmission (ii) deletions in the Spike protein NTD map to an antigenic supersite, (iii) the repertoire of deletions in the Spike protein is expanding over the course of the pandemic and (iv) deletions are present in a subset of vaccine breakthrough variants. Indeed, deletion mutations are not operating independent of other mutation classes. In addition to deletion mutations, several substitution mutations are also associated with surges in cases (e.g. L452R and T478K in the receptor binding domain; **Figure S1**). Thus, a concerted evolution of strategically placed deletions and substitutions appears to be conferring SARS-CoV-2 with the fitness to evade immunity and achieve efficient transmission between hosts (**Figure 6**).

**Figure 6.**
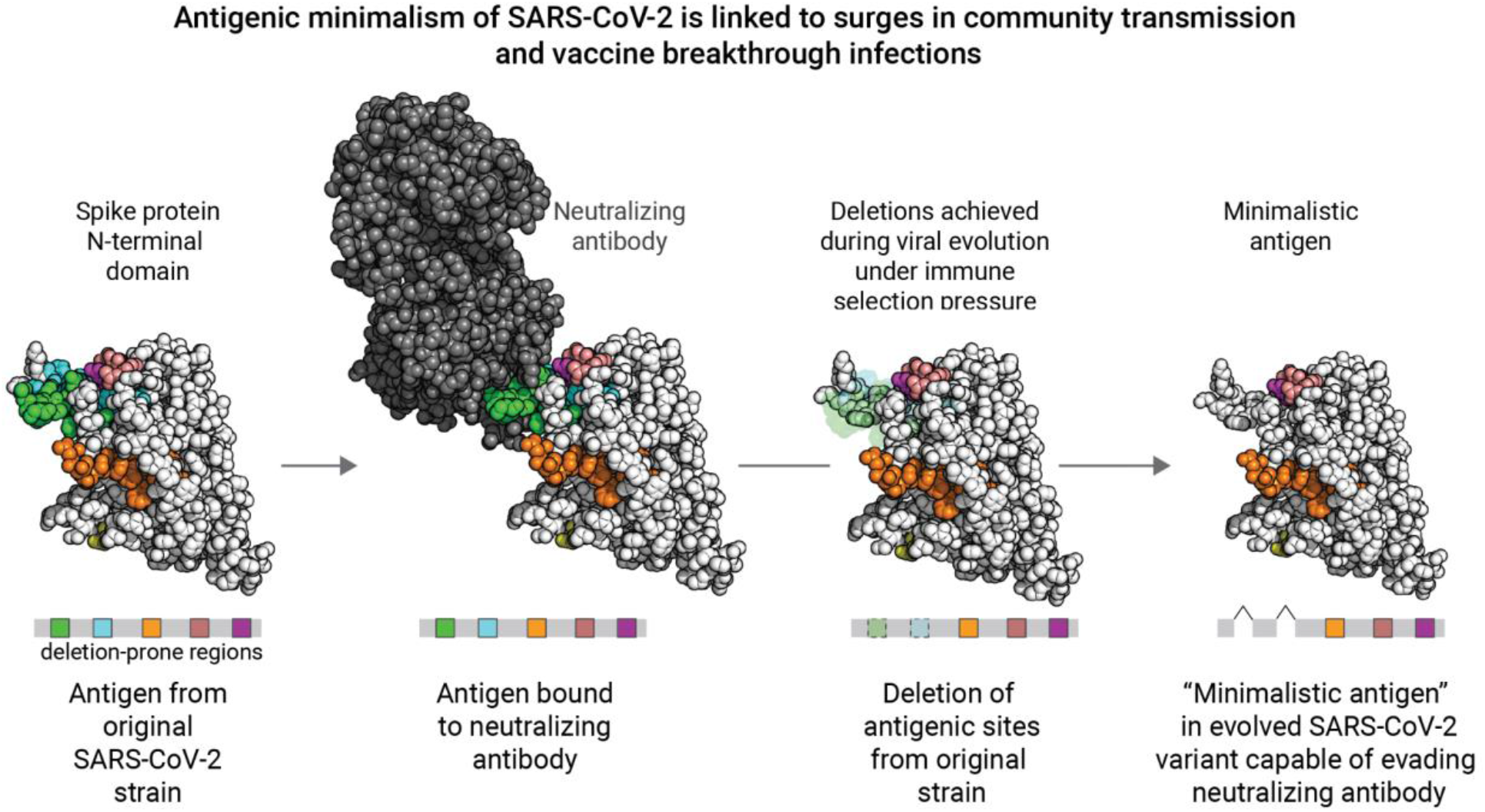
Schematic overview of the Spike protein N-terminal domain acquiring deletion mutations during evolution to evade immune response. The deletion mutations occur concurrently with other substitution mutations (not highlighted) in the background.

Our finding that Spike protein NTD deletions are strongly enriched for association with test positivity surges is notable in the context of a previous report identifying the NTD as the most common site of deletions^15^. Specifically, this prior study highlighted four recurrent deletion regions in the NTD based on the GISAID data deposited as of October 2020 (146,795 total sequences). Several of these regions overlap with the residues of the recently identified NTD antigenic supersite, and deletions within them can abrogate binding to neutralizing antibodies^13–15^. Our findings build upon this prior work by examining the deletions which have arisen in the interim, during which over 1.1 million additional sequences have been deposited. In addition to validating the previously suggested definitions of recurrent deletion regions RDR1 (ΔH69/V70 and flanking deletions), RDR2 (ΔY144 and flanking deletions), and RDR3 (ΔI210 and ΔN211), we found that RDR4 (previously defined as positions 242-248) has recently expanded to include positions 249-253. These residues are indeed part of the structurally mapped supersite^13,14^, and a variant with the Δ246-253 deletion increased in prevalence during a recent test positivity surge in Chile. The recently evolved ΔF157/R158 deletion, which has expanded during the massive surge in India, marks a new recurrent deletion region which also maps to the supersite^14^. Finally, our real time surveillance of clinically annotated SARS-CoV-2 genomes among COVID-19 cases at the Mayo Clinic, including vaccine breakthrough infections, revealed contiguous deletions (Δ85-90 and Δ167-174) that have not been recognized as recurrent deletion regions previously. The proximity of these regions to the antigenic supersite suggests that they may become more prevalent and that deletions in these regions should be monitored for associations with future surges. The striking trend that the most frequently deleted NTD regions are proximal to a single antigenic supersite highlights the prominent role that host immunity has played in shaping the genomic evolution of SARS-CoV-2 from the beginning of this pandemic.

There are a few limitations of this study. First, the geographic distribution of sequences deposited in the GISAID database is not representative of the global population, with a majority of the sequences coming from the United States or the United Kingdom. Future genomic epidemiology studies would be improved by expanded sequencing efforts in other countries. Second, the identification of mutations associated with surges during early months of the pandemic is complicated by the relative paucity of whole genome sequencing data deposited during that time. Third, the publicly accessible genomic data is generally not linked to relevant phenotypic information (e.g., disease severity) or relevant medical histories (e.g., comorbidities and vaccination status). Thus, while we are able to identify correlations between mutational prevalence and case surges, we cannot determine whether particular mutations are associated with more severe disease or are observed more frequently than expected by chance in vaccinated individuals. While the latter shortcoming is partially addressed by our independent whole genome sequencing of virus isolated from COVID-19 cases with accessible longitudinal records (including previously vaccinated individuals), this analysis was limited by the small size of the cohort (n = 53) and the lack of corresponding antibody titer data. We plan to perform sequencing of more SARS-CoV-2 samples from COVID-19 patients.

Taken together, by synthesizing insights from genomic epidemiology and clinical genomics, we have uncovered that SARS-CoV-2 likely employs antigenic minimalism in the Spike protein as a strategy to evade immune responses induced by infection or vaccination. These findings have important therapeutic and public health policy implications. The repertoire of deletion mutations in the N-terminal domain should be considered when developing future vaccines and biologics to counter the immuno-evasive strategies of SARS-CoV-2. From the public health standpoint, we must expand sequencing efforts around the world and encourage the transparent linking of relevant deidentified patient phenotypic data (e.g. disease severity, vaccination status) to each deposited SARS-CoV-2 genome. While the current analysis focuses on the Spike protein, future work focusing on other SARS-CoV-2 proteins, such as the nucleocapsid protein and RNA-directed RNA polymerase, will shed light on the role of the overall mutational landscape of the SARS-CoV-2 proteome for viral fitness and immune evasion. Such a holistic understanding of the mutational landscape of SARS-CoV-2 is imperative to proactively predict variants that could trigger outbreaks and vaccine breakthroughs, as well as guide the development of comprehensive therapeutic strategies to defeat the COVID-19 pandemic.

## Methods

### Analysis of publicly deposited SARS-CoV-2 genomic sequences

Around 1.57 million SARS-CoV-2 genome sequences (with 1,285 unique lineages and 10,298 unique spike mutations) were obtained from GISAID^17^ (data retrieved from https://www.gisaid.org/ as of 18th May 2021) across 187 countries/territories. Using the Wuhan-Hu-1 sequence as reference (UniProt ID: P0DTC2), there have been 10,298 amino acid mutations in the Spike protein detected in at least one sequence. To filter out potential sequencing artifacts, we excluded mutations that were present in fewer than 100 sequences, resulting in 857 unique Spike protein mutations.

### Identification of surge-associated SARS-CoV-2 mutations

To identify mutations that have been temporally associated with surges in COVID-19 cases throughout the pandemic, we assessed monthly mutational prevalences and test positivity over three-month intervals in each country. For each of the 857 mutations, the monthly mutational prevalence was computed for a given country as:

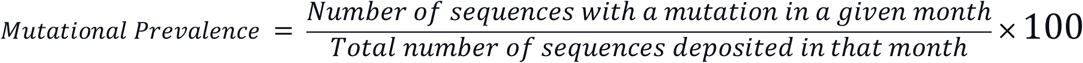

Positivity data for PCR tests was obtained from the OWID resource^28,29^ (retrieved from https://github.com/owid/covid-19-data/tree/master/public/data on May 21, 2021). For each country, the monthly test positivity was calculated as:

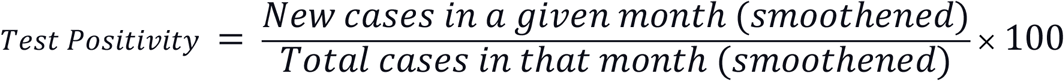

To identify surge-associated mutations, we classified the monthly mutational prevalence (for each mutation) and the monthly test positivity as increasing (monotonically), decreasing (monotonically), or mixed over sliding three-month intervals over the course of the pandemic. Any mutation which monotonically increased in prevalence over this interval in a country with a simultaneous monotonic increase in test positivity was defined as a “surge-associated mutation.” There were 89 such mutations.

### Comparison of surge-associated mutations to mutations in CDC variants of interest and concern

In order to test the value of our method, we obtained the set of CDC variants of interest and concern as of April 15, 2021^8^. At this time, there were 5 variants of concern and 8 variants of interest, with no variants of high consequence. From the 13 classified variants, there 56 unique mutations listed, of which 25 were found only in variants of interest, 24 were found only in variants of concern, and 7 were found in both variants of interest and concern. After identifying the surge-associated mutations as described above, we determined the fraction of mutations comprising the CDC-classified variants which were captured by this approach.

### Assessment of mutation types for enrichment of surge-associated mutations

After identifying the 89 surge-associated mutations, we tested whether any of the contributing mutation types (deletions, insertions, or substitutions) were enriched for surge-associated mutations. To do so, we constructed a 2 × 3 table giving the number of surge-associated and non-surge-associated mutations in each category. To determine whether one or more groups showed a statistically significant enrichment, a chi-square p-value was calculated using the chisq.test function from the stats package (4.0.3) in R. Post-hoc tests were performed by considered constructing 2×2 contingency tables to compare each mutation type against all others. Then, odds ratios and their corresponding 95% confidence intervals were calculated using the fisher.test function from the stats package (version 4.0.3) in R.

### Identification of new recurrent deletion regions in the Spike protein

Recurrent deletion regions (RDRs) were previously defined as four sites within the NTD to which over 90% of all Spike protein deletions occurred, per the 146,795 SARS-CoV-2 sequences deposited in GISAID as of October 24, 2020. To identify potential new RDRs that have emerged since this time, we first plotted the distribution of deletion counts for each amino acid (i.e. number of sequences in which deletion of the given amino acid was observed) in the Spike protein, considering all 1,313,962 sequences analyzed as of April 30, 2021. We calculated the 95th percentile of the deletion count distribution, which is 22.4. We then bucketed each residue *R* into categories (Yes, No, Possible) reflecting whether or not it should be considered as part of an RDR (i.e, a contiguous stretch of two or more amino acid residues which undergo deletion events more frequently than expected by chance) as follows (illustrated schematically in **Table S4**).

Once each residue was categorized in this way, then any residue *P* in the “Possible” category were subjected to further analysis to convert their labels into “Yes” or “No.” Specifically, we took a step-wise approach, walking in both directions from *P* until the first encounter of a residue categorized as “Yes” or “No” (i.e., other residues labeled as “Possible” were ignored). If a residue categorized as “Yes” was encountered before any residue categorized as “No” in either direction, then the “Possible” label was converted to “Yes.” If a residue categorized as “No” was encountered before any residue categorized as “Yes” in both directions, then the “Possible” label was converted to “Yes.”

With each residue categorized as “Yes” or “No”, we then simply merged the residue windows with consecutive “Yes” labels to define the updated set of Spike protein RDRs. We name the RDRs on the basis of the first and last amino acid residues contained within the region; for example, the RDR including residues C14, Q15, and V16 is defined as RDR_14-16_.

### Temporal analysis of expansions in recurrent deletion regions

To assess the expansion of regions undergoing deletions over time, we plotted a time series heatmap indicating the first time (month) at which a given deletion was identified across all GISAID sequences, and the number of sequences in which that deletion was detected in that month and all subsequent months. The residues plotted were defined based on the definition of RDRs provided above, which builds upon the regions defined previously ^15^.

### Structural analysis of SARS-CoV-2 Spike protein

Structural analyses and illustrations were performed in PyMOL (version 2.3.4). The cryo-EM structure of the Spike protein characterizing the interaction with a neutralizing antibody 4A8 (PDB identifier: 7C2L), described by Chi et al.^21^, was retrieved from the PDB.

### Amplicon sequencing of SARS-CoV-2 genome obtained from individuals with breakthrough infections

This is a retrospective study of individuals who underwent polymerase chain reaction (PCR) testing for suspected SARS-CoV-2 infection at the Mayo Clinic and hospitals affiliated to the Mayo health system. This study was reviewed by the Mayo Clinic Institutional Review Board and determined to be exempt from human subjects research. Subjects were excluded if they did not have a research authorization on file.

SARS-CoV-2 RNA-positive upper respiratory tract swab specimens from patients with vaccine breakthrough or reinfection of COVID-19 were subjected to next-generation sequencing, using the commercially available Ion AmpliSeq SARS-CoV-2 Research Panel (Life Technologies Corp., South San Francisco, CA) based on the “sequencing by synthesis” method. The assay amplifies 237 sequences ranging from 125 to 275 base pairs in length, covering 99% of the SARS-CoV-2 genome. Viral RNA was first manually extracted and purified from these clinical specimens using MagMAX™ Viral / Pathogen Nucleic Acid Isolation Kit (Life Technologies Corp.), followed by automated reverse transcription-PCR (RT-PCR) of viral sequences, DNA library preparation (including enzymatic shearing, adapter ligation, purification, normalization), DNA template preparation, and sequencing on the automated Genexus™ Integrated Sequencer (Life Technologies Corp.) with the Genexus™ Software version 6.2.1. A no-template control and a positive SARS-CoV-2 control were included in each assay run for quality control purposes. Viral sequence data were assembled using the Iterative Refinement Meta-Assembler (IRMA) application (50% base substitution frequency threshold) to generate unamended plurality consensus sequences for analysis with the latest versions of the web-based application tools: Pangolin^30^ for SARS-CoV-2 lineage assignment; Nextclade^31^ for viral clade assignment, phylogenetic analysis, and S codon mutation calling, in comparison to the wild-type reference sequence of SARS-CoV-2 Wuhan-Hu-1 (lineage B, clade 19A).

## Data Availability

After publication, the data will be made available upon reasonable requests to the corresponding author. A proposal with detailed description of study objectives and the statistical analysis plan will be needed for evaluation of the reasonability of requests. Deidentified data will be provided after approval from the corresponding author and the Mayo Clinic.

## Acknowledgements

The authors thank Murali Aravamudan and Eric Schadt for the careful review and feedback on this manuscript.

## Author Contributions

VS conceived the study. AJV and VS advanced the study design and generated the hypotheses. PA, PL, PG, AJV and VS wrote the manuscript and reviewed the findings. AJV, PA, PL, PG, RS, AS, DRC, JDY, and JCO contributed methods, data, analysis, or software. JCOH, JDY, BSP, AN, RTH, ADB, and JDH reviewed the study, findings, and the manuscript. All authors revised the manuscript.

## Supplementary Information

**Figure S1.**
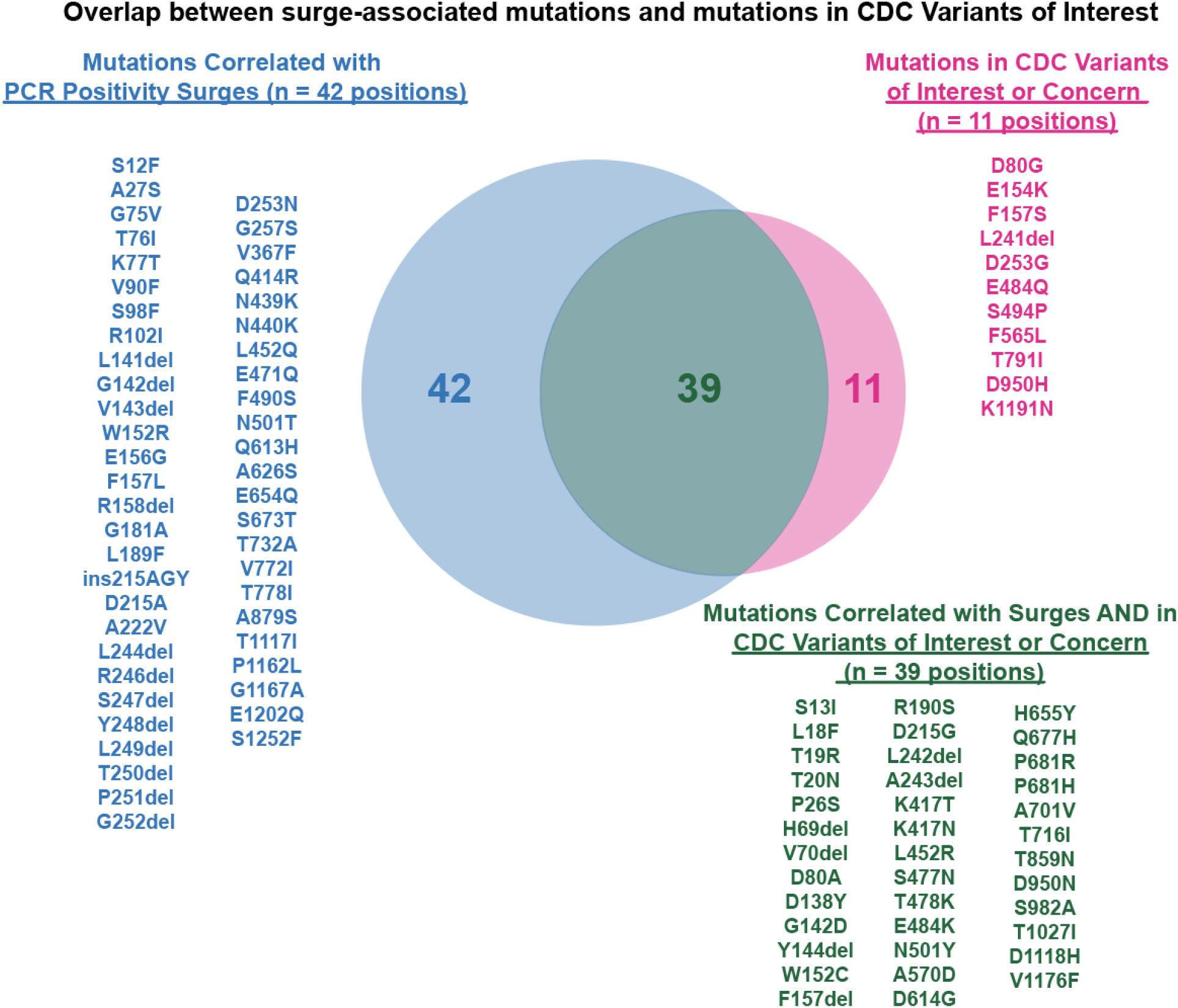
Comparison of surge-associated mutations identified in this study and mutations present in variants of interest or concern as categorized by the Centers for Disease Control & Prevention.

**Figure S2.**
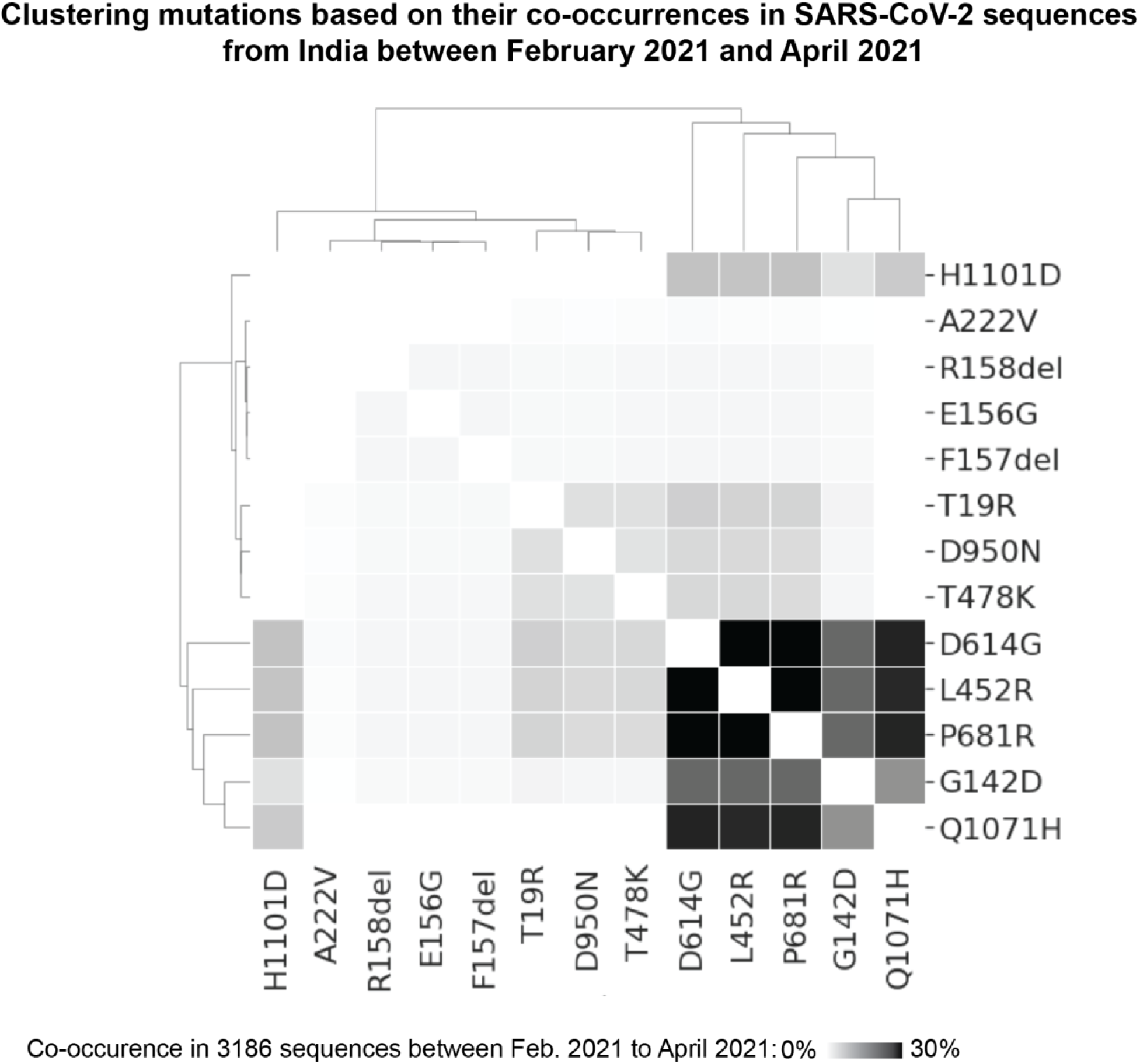
Clustering mutations based on their co-occurrences in SARS-CoV-2 sequences from India between February 2021 and April 2021.

**Table S1:** List of GISAID accession IDs with the same recurrent deletions observed as seen in the vaccine breakthrough patients.

| Accession ID | Location | Collection date | RDR Stretch | Deletions |
| --- | --- | --- | --- | --- |
| EPI_ISL_1286100 | Europe / Germany / Hamburg | 2021-03-06 | 85-90 | ΔP85;ΔF86;ΔN87;ΔD88;ΔG89;ΔV90 |
| EPI_ISL_1725555 | Europe / Germany / Berlin | 2021-03-10 | 85-90 | ΔP85;ΔF86;ΔN87;ΔD88;ΔG89;ΔV90 |
| EPI_ISL_1840693 | North America / USA / Texas | 2021-04-17 | 85-90 | ΔP85;ΔF86;ΔN87;ΔD88;ΔG89;ΔV90 |
| EPI_ISL_741037 | Europe / United Kingdom / England | 2020-11-12 | 156-164 | ΔE156;ΔF157;ΔR158;ΔV159;ΔY160;ΔS161;ΔS162;ΔA163;ΔN164 |
| EPI_ISL_1389681 | Europe / United Kingdom / Scotland | 2021-03-19 | 156-164 | ΔE156;ΔF157;ΔR158;ΔV159;ΔY160;ΔS161;ΔS162;ΔA163;ΔN164 |
| EPI_ISL_1321491 | South America / Chile / Region Metropolitana de Santiago / La Florida | 2021-02-11 | 156-164 | ΔE156;ΔF157;ΔR158;ΔV159;ΔY160;ΔS161;ΔS162;ΔA163;ΔN164 |
| EPI_ISL_1841239 | Asia / India / Gujarat / Ahmedabad | 2021-04-18 | 167-174 | ΔT167;ΔF168;ΔE169;ΔY170;ΔV171;ΔS172;ΔQ173;ΔP174 |
| EPI_ISL_1703668 | Asia / India / Maharashtra | 2021-03-11 | 167-174 | ΔT167I;ΔF168;ΔE169;ΔY170;ΔV171I;ΔS172;ΔQ173;ΔP174 |

**Table S2:** All the mutations in the spike protein that have positive correlation with the test positivity percentage across the recent timeline (Feb-Apr 2021) of pandemic in India has been tabulated here. Following are the expansion of the abbreviations used in the table header - Total Seqs. Dep. : Total number of sequences deposited in the particular month in India. Test Pos.% : Test positivity percentage, Mut Prev.%: Mutation prevalence percentage, Rho (Pearson) Mut Prev.% vs Test Pos.% : The Pearson correlation Rho value between test positivity and mutational prevalence, Test pos. List : test positivity percentage over the window of 3 months (February 2021 to April 2021), Mut Prev. List : mutation prevalence percentage over the window of 3 months, MaxΔ Mut Prev. : maximum difference in the mutational prevalence percentage observed over the window of 3 months. Total sequences deposited.

| **Spike Mutation** | **Test Positivity %** | **Mutation Prevalence %** | **Rho (Pearson) Mutational Prevalence.% vs. Test Positivity.%** | **Test Positivity. List (3 months)** | **Mutational Prevalence List (3 months)** | **Max Δ Mut Prev. (3 months)** |
| --- | --- | --- | --- | --- | --- | --- |
| D614G | 11.276 | 100 | 0.823 | (1.76, 2.896, 11.276) | (99.043, 99.681, 100.0) | 0.957 |
| A222V | 11.276 | 8.174 | 1 | (1.76, 2.896, 11.276) | (0.319, 1.214, 8.174) | 7.855 |
| L452R | 11.276 | 68.392 | 0.925 | (1.76, 2.896, 11.276) | (26.954, 46.773, 68.392) | 41.438 |
| T478K | 11.276 | 29.155 | 0.99 | (1.76, 2.896, 11.276) | (0.239, 7.668, 29.155) | 28.916 |
| P681R | 11.276 | 68.937 | 0.953 | (1.76, 2.896, 11.276) | (26.236, 43.642, 68.937) | 42.701 |
| G142D | 11.276 | 23.433 | 0.615 | (1.76, 2.896, 11.276) | (12.281, 23.067, 23.433) | 11.152 |
| Q1071H | 11.276 | 31.608 | 0.72 | (1.76, 2.896, 11.276) | (17.225, 29.01, 31.608) | 14.383 |
| D950N | 11.276 | 25.613 | 0.99 | (1.76, 2.896, 11.276) | (1.116, 7.476, 25.613) | 24.497 |
| T19R | 11.276 | 30.79 | 0.991 | (1.76, 2.896, 11.276) | (1.116, 8.498, 30.79) | 29.674 |
| ΔR158 | 11.276 | 14.986 | 0.997 | (1.76, 2.896, 11.276) | (1.116, 1.534, 14.986) | 13.87 |
| ΔF157I | 11.276 | 14.986 | 0.997 | (1.76, 2.896, 11.276) | (1.116, 1.534, 14.986) | 13.87 |
| E156G | 11.276 | 14.986 | 0.997 | (1.76, 2.896, 11.276) | (1.116, 1.534, 14.986) | 13.87 |
| H1101D | 11.276 | 14.441 | 0.845 | (1.76, 2.896, 11.276) | (6.459, 11.502, 14.441) | 7.982 |

**Table S3:** All the mutations in the spike protein that have positive correlation with the test positivity percentage across the recent timeline of pandemic (Feb-Apr 2021) in Chile has been tabulated here. Following are the expansion of the abbreviations used in the table header - Total Seqs. Dep. : Total number of sequences deposited in the particular month in Chile. Test Pos.% : Test positivity percentage, Mut Prev.%: Mutation prevalence percentage, Rho (Pearson) Mut Prev.% vs Test Pos.% : The Pearson correlation Rho value between test positivity and mutational prevalence, Test pos. List : test positivity percentage over the window of 3 months, Mut Prev. List : mutation prevalence percentage over the window of 3 months, MaxΔ Mut Prev. : maximum difference in the mutational prevalence percentage observed over the window of 3 months.

| **Spike Mutation** | **Test Positivity %** | **Mutation Prevalence %** | **Rho (Pearson) Mutational Prevalence.% vs. Test Positivity.%** | **Test Pos. List (3 months)** | **Mut Prev. List (3 months)** | **Max Δ Mut Prev. (3 months)** |
| --- | --- | --- | --- | --- | --- | --- |
| L18F | 11.218 | 31.409 | 0.954 | (7.265, 8.632, 11.218) | (4.274, 23.51, 35.683) | 31.409 |
| T20N | 11.218 | 32.238 | 0.965 | (7.265, 8.632, 11.218) | (2.564, 21.192, 34.802) | 32.238 |
| P26S | 11.218 | 32.678 | 0.944 | (7.265, 8.632, 11.218) | (2.564, 23.51, 35.242) | 32.678 |
| ΔI68 | 11.218 | 4.431 | 0.948 | (7.265, 8.632, 11.218) | (0.855, 3.642, 5.286) | 4.431 |
| ΔH69 | 11.218 | 7.699 | 0.978 | (7.265, 8.632, 11.218) | (6.838, 10.927, 14.537) | 7.699 |
| ΔV70 | 11.218 | 7.699 | 0.978 | (7.265, 8.632, 11.218) | (6.838, 10.927, 14.537) | 7.699 |
| ΔS71 | 11.218 | 4.431 | 0.948 | (7.265, 8.632, 11.218) | (0.855, 3.642, 5.286) | 4.431 |
| ΔG72 | 11.218 | 4.431 | 0.948 | (7.265, 8.632, 11.218) | (0.855, 3.642, 5.286) | 4.431 |
| ΔT73 | 11.218 | 4.431 | 0.948 | (7.265, 8.632, 11.218) | (0.855, 3.642, 5.286) | 4.431 |
| G75V | 11.218 | 21.796 | 0.803 | (7.265, 8.632, 11.218) | (6.838, 27.152, 28.634) | 21.796 |
| T76I | 11.218 | 23.092 | 0.803 | (7.265, 8.632, 11.218) | (5.983, 27.483, 29.075) | 23.092 |
| D138Y | 11.218 | 31.797 | 0.941 | (7.265, 8.632, 11.218) | (2.564, 23.179, 34.361) | 31.797 |
| ΔY144 | 11.218 | 1.559 | 0.981 | (7.265, 8.632, 11.218) | (7.692, 7.947, 9.251) | 1.559 |
| R190S | 11.218 | 32.678 | 0.947 | (7.265, 8.632, 11.218) | (2.564, 23.179, 35.242) | 32.678 |
| ΔR246 | 11.218 | 26.202 | 0.812 | (7.265, 8.632, 11.218) | (6.838, 30.795, 33.04) | 26.202 |
| ΔS247 | 11.218 | 26.202 | 0.812 | (7.265, 8.632, 11.218) | (6.838, 30.795, 33.04) | 26.202 |
| ΔY248 | 11.218 | 26.202 | 0.812 | (7.265, 8.632, 11.218) | (6.838, 30.795, 33.04) | 26.202 |
| ΔL249 | 11.218 | 26.202 | 0.812 | (7.265, 8.632, 11.218) | (6.838, 30.795, 33.04) | 26.202 |
| ΔT250 | 11.218 | 26.202 | 0.812 | (7.265, 8.632, 11.218) | (6.838, 30.795, 33.04) | 26.202 |
| ΔP251 | 11.218 | 26.202 | 0.812 | (7.265, 8.632, 11.218) | (6.838, 30.795, 33.04) | 26.202 |
| ΔG252 | 11.218 | 26.202 | 0.812 | (7.265, 8.632, 11.218) | (6.838, 30.795, 33.04) | 26.202 |
| D253N | 11.218 | 26.202 | 0.812 | (7.265, 8.632, 11.218) | (6.838, 30.795, 33.04) | 26.202 |
| K417T | 11.218 | 20.743 | 1 | (7.19, 8.632, 11.218) | (1.724, 9.603, 22.467) | 20.743 |
| L452Q | 11.218 | 26.669 | 0.818 | (7.265, 8.632, 11.218) | (7.692, 31.788, 34.361) | 26.669 |
| E484K | 11.218 | 30.14 | 0.933 | (7.265, 8.632, 11.218) | (5.983, 26.159, 36.123) | 30.14 |
| F490S | 11.218 | 27.523 | 0.83 | (7.265, 8.632, 11.218) | (6.838, 31.126, 34.361) | 27.523 |
| N501Y | 11.218 | 35.946 | 0.934 | (7.265, 8.632, 11.218) | (8.547, 32.45, 44.493) | 35.946 |
| A570D | 11.218 | 3.268 | 0.985 | (7.265, 8.632, 11.218) | (5.983, 7.616, 9.251) | 3.268 |
| H655Y | 11.218 | 28.819 | 0.92 | (7.265, 8.632, 11.218) | (5.983, 26.159, 34.802) | 28.819 |
| P681H | 11.218 | 0.29 | 0.923 | (7.265, 8.632, 11.218) | (9.402, 9.603, 9.692) | 0.29 |
| T716I | 11.218 | 2.413 | 0.836 | (7.265, 8.632, 11.218) | (6.838, 8.94, 9.251) | 2.413 |
| T859N | 11.218 | 27.523 | 0.83 | (7.265, 8.632, 11.218) | (6.838, 31.126, 34.361) | 27.523 |
| S982A | 11.218 | 3.268 | 0.958 | (7.265, 8.632, 11.218) | (5.983, 7.947, 9.251) | 3.268 |
| T1027I | 11.218 | 33.559 | 0.939 | (7.265, 8.632, 11.218) | (2.564, 24.503, 36.123) | 33.559 |
| D1118H | 11.218 | 3.268 | 0.958 | (7.265, 8.632, 11.218) | (5.983, 7.947, 9.251) | 3.268 |
| V1176F | 11.218 | 29.259 | 0.924 | (7.265, 8.632, 11.218) | (5.983, 26.159, 35.242) | 29.259 |

**Table S4:** Schematic representation of the decision schema for considering a residue R to be a part of a RDR. Deletion count of <=22 is represented by ✗. Deletion count of >=23 is represented by ✔.

| Deletion count |  |  |  |  | Should Residue *R* be considered part of a RDR? |
| --- | --- | --- | --- | --- | --- |
| *R* - 2 | *R* - 1 | Residue *R* | *R* + 1 | *R* + 2 |  |
|  | X | X |  |  | No |
|  |  | X | X |  | No |
|  | ✓ | ✓ |  |  | Yes |
|  |  | ✓ | ✓ |  | Yes |
|  | ✓ |  | ✓ | ✓ | Yes |
| ✓ | ✓ |  | ✓ |  | Yes |
| X | X | ✓ |  | ✓ | Possible |
| ✓ |  | ✓ | X | X | Possible |
|  | ✓ | X | ✓ |  | Possible |

